# Longitudinal, Multicenter Study of Clinical Factors Impacting Health-Related Quality of Life in Pediatric Autoimmune Liver Disease

**DOI:** 10.1101/2025.05.19.25327910

**Authors:** Rebecca Farr, Cyd Castro Rojas, Mosab Alquraish, Kevin Hommel, Rashmi Sahay, Alexander Weymann, Katelyn Saarela, Sakil Kulkarni, Mary Ayers, James Squires, Amy Taylor, Heidi J. Kalkwarf, Alexander Miethke

## Abstract

**Background:** Children with autoimmune liver disease (AILD) face unique challenges that may impair their health-related quality of life (HRQoL). This multicenter, prospective, longitudinal study evaluated HRQoL over time and identified associated clinical factors.

**Methods:** A total of 162 participants from five centers completed at least one HRQoL assessment. Medical and laboratory data were abstracted within three months of each assessment. Fatigue and pruritus were reported at one center. Generalized linear mixed-effects modeling was used to examine longitudinal associations between HRQoL and clinical variables.

**Results:** Participants reported the lowest HRQoL scores in school and emotional domains, while the physical and social domains were less affected. Compared to healthy children, participants with AILD reported lower overall HRQoL. Longitudinal analysis revealed that caregivers of participants with overlap syndrome reported higher emotional, social, psychosocial, and total scores. Paradoxically, children with disease complications had better school scores, possibly due to increased support services. Prednisone use was associated with improved emotional scores, while azathioprine use was associated with lower social scores. Elevated ALT levels were associated with lower HRQoL scores, particularly when reported by caregivers. Disease duration and presence of inflammatory bowel disease were not significantly associated with HRQoL.

From one center, fatigue and pruritus were significantly associated with lower HRQoL, especially in the physical, psychosocial, and total domains. Fatigue was also associated with elevated liver enzymes and reduced rates of biochemical remission, suggesting that inflammation may contribute to a disrupted liver-brain axis.

**Conclusion:** These findings underscore the multidimensional impact of AILD on pediatric patients and highlight the need for further research into the pathophysiology of fatigue and potential therapeutic interventions.

## Introduction

Pediatric autoimmune liver disease (AILD) is a spectrum of rare and severe chronic liver diseases, ranging from autoimmune hepatitis (AIH) without biliary tract involvement to primary sclerosing cholangitis (PSC) with injury to the small and/or large bile ducts. Patients with autoimmune sclerosing cholangitis (ASC) display signs and symptoms of AIH and PSC ^1,2^. Children and young adults with AILD face disabling symptoms associated with their diseases, and while AILD itself is often managed with a combination of corticosteroids and immunosuppressants, options for symptom-specific relief, such as for severe fatigue or pruritus, are limited ^2,3^. Additionally, these patients are at risk for progressive cirrhosis, sometimes necessitating life-altering liver transplantation ^4–6^. As a result of these factors, the health-related quality of life (HRQoL) of this patient population may be impaired.

To date, research on HRQoL in pediatric AILD is limited to single centers and examines HRQoL of patients at only one point in time. Gulanti et al. performed a cross-sectional study of 30 patients with AILD who completed a single HRQoL survey an average of 4.6 years after diagnosis and showed impairment in quality of life compared to healthy controls ^7^. A subsequent cross-sectional study by Bozzini et al. examined 80 patients with AIH specifically and discovered that patients treated with a lower prednisone dose had higher physical well-being scores at time of interview ^8^. Lastly, Trevizoli et al. performed a cross-sectional study of 43 participants with AIH and found that daily use of medication led to lower psychosocial scores ^9^.

Literature evaluating HRQoL in adults with chronic liver disease has shown that factors negatively impacting HRQoL are often dependent on the underlying etiology of liver disease ^10^. Fatigue has emerged as a key driver of reduced HRQoL, especially among patients with autoimmune liver diseases ^11^. Notably, a recent prospective multicenter study of adults with autoimmune hepatitis (AIH) found that physical HRQoL was significantly impaired compared to healthy controls, and that HRQoL impairment was closely associated with the presence of fatigue as well as symptoms of depression and anxiety ^12^.

To our knowledge, there are no published longitudinal, multicenter studies examining HRQoL in children and young adults with AILD. Therefore, our aim was to better understand the variability of HRQoL at multiple timepoints and at several institutions and to identify clinical factors that may influence quality of life in this patient population.

## Methods

### Study design and participants

We performed a longitudinal, prospective cohort study that included participants with AILD from three institutional review board-approved studies. The studies are conducted in accordance with the Declaration of Helsinki. Two of the studies were single center studies conducted at Cincinnati Children’s Hospital Medical Center (CCHMC, Cincinnati, OH) and have been on-going since 2016 (NCT03178630; IRB ID: 2016-7388) and 2017 (IRB ID: 2017-2284). The third study was a collaborative, multi-center learning health network, Autoimmune Liver disease Network for Kids (A-LiNK, NCT05750498, IRB ID: 2020-1019) that was established in 2021. A total of five sites in A-LiNK were included in this study: CCHMC, Nationwide Children’s Hospital (Columbus, OH), Seattle Children’s Hospital (Seattle, WA), St. Louis Children’s Hospital - Washington University (St Louis, MO), and University of Pittsburgh Medical Center Children’s Hospital of Pittsburgh (Pittsburgh, PA). All centers obtained IRB approval by relying on a single protocol at CCHMC (Cincinnati Children’s Hospital IRB, FWA #00002988). If a participant was enrolled in a CCHMC single-center study, as well as A-LiNK, the participant was only included once, and the participant’s information from all three studies was combined under one record entry.

We included children and young adults with AIH, PSC or AIH/PSC overlap syndrome who were between the ages of 1-25 years and had completed at least one validated pediatric assessment of quality of life, Pediatric Quality of Life Inventory™ Version 4.0 (PedsQL 4.0) Core Short Form ^13,14^. Enrolled participants met the clinical definition of AIH, PSC or AIH/PSC overlap syndrome based on the AASLD disease-specific guidelines ^4,15^. Participants listed for or with a history of liver transplantation were excluded.

An explanation of the study was given to participants who met eligibility criteria, and participants or their legal guardians provided written informed consent/assent.

### Data collection

Quality of life was assessed with the validated PedsQL 4.0 Generic Core Short Form, which has developmentally appropriate assessments by age. The 15-item PedsQL 4.0 Generic Core Short Form includes questions regarding the following four domains: 1) Physical Functioning (5 items), 2) Emotional Functioning (4 items), 3) Social Functioning (3 items), and 4) School Functioning (3 items). Participants and their caregivers separately rated the degree to which the item had been an issue for them during the past month, ranging from 0 (never) to 4 (almost always). Items were reverse-scored and linearly transformed to a 0 to 100 scale (higher scores reflect better HRQoL). The psychosocial domain score was obtained by averaging the items from the emotional, social and school domains. The total domain score was obtained by averaging the items from all four domains. The surveys were administered using two methods, paper surveys during the clinical care process or via email after the visit. The method of survey collection has previously been shown not to impact results ^14^. If a participant completed the PedsQL on multiple occasions, each response was recorded (up to a maximum of 3), as long as there was at least a 6-month time period between each form.

Additionally, we obtained data from the electronic medical record for the CCHMC cohort studies and from the A-LiNK registry for the multicenter A-LiNK cohort. Collected data included participant demographics, date and diagnosis of AILD (AIH, PSC, AIH/PSC overlap syndrome), disease-specific factors, and laboratory values. Only disease-specific factors and labs within 3 months of each PedsQL 4.0 form were used. The factors included presence of concurrent inflammatory bowel disease (IBD), number of AILD complications (i.e., esophageal variceal bleed, ascites, hepatic encephalopathy, and cholangitis), and use of prednisone and azathioprine. Laboratory measures included alanine aminotransferase (ALT), aspartate aminotransferase (AST), alkaline phosphatase (Alk Phos), gamma-glutamyl transpeptidase (GGT), total bilirubin, total protein, immunoglobulin G (IgG), platelets and International Normalized Ratio (INR). AST to platelet ratio index (APRI) and Fibrosis-4 (FIB-4) were calculated using the abstracted demographics and laboratory values ^16,17^.

Symptoms of fatigue and pruritus were obtained exclusively from the CCHMC single-center cohort and were included only if documented within 3 months of each HRQoL form. Presence of fatigue was assessed either by a research survey or from clinic notes, while pruritus was assessed solely by clinic notes. Data were recorded as "yes," "no," or "unknown." Notably, fatigue and pruritus were coded as present only if explicitly mentioned in the clinic notes.

Data were collected in a Center for Clinical & Translational Science & Training (CCTST) Research Electronic Data Capture (REDCap) database, with quality control according to proper Standard Operating Procedures.

### Statistical analysis

Descriptive statistics were used to summarize all demographic, clinical, and laboratory measurements. We compared participant demographics, disease characteristics, and baseline HRQoL scores between CCHMC and the other institutions by a Chi-square or Fisher’s exact test for categorical data and a Wilcoxon rank-sum test for continuous data. Pearson correlation was used to assess the relationship between the HRQoL baseline scores of participants and caregivers.

We evaluated the longitudinal association between repeated HRQoL assessments and clinical factors and labs using a generalized linear mixed-effects model. Study participants were treated as a random effect to account for the non-independence of observations within individuals. First, we assessed the HRQoL scores across different time points. Next, we fitted models to evaluate the effects of assessment time point and clinical factors, as well as their interaction, on HRQoL scores. These models were fitted separately for each clinical factor. We additionally fitted models to evaluate the effects of participant biochemistries (AST, ALT, Alk phos, and GGT) and assessment time points. Biochemistries were log-transformed to normalize their skewed distributions. Finally, we fitted multivariable longitudinal models including all clinical factors to account for potential collinearity between factors for each HRQoL score. Statistical significance was considered if p ≤ 0.05 in all models.

Additionally, we used the concept of minimal clinically important difference (MCID) to determine which score changes were clinically significant. MCID is defined as the smallest difference in scores that patients perceive as meaningful and is dependent on the HRQoL assessment tool used. For PedsQL, the MCID was derived from the standard error of measurement in a large population-based study and is defined as 4.4 points for child self-report and 4.5 points for caregiver-proxy report ^14^.

## Results

In the single-center CCHMC studies, 154 individuals were screened for eligibility, of whom 63 were excluded because they did not have AILD or had incomplete/missing PedsQL forms (Figure 1). From the A-LiNK study, 137 individuals were screened for eligibility and 66 were excluded due to incomplete/missing PedsQL forms or missing clinical information. In the CCHMC cohort, a significantly higher number of males were excluded due to missing or incomplete PedsQL forms (Supplemental Table 1). After screening, a total of 162 participants were included in this study. Broken down by site, 104 participants were from CCHMC, 26 from Nationwide Children’s Hospital, 6 from Seattle Children’s Hospital, 5 from St. Louis Children’s Hospital - Washington University, and 21 from University of Pittsburgh Medical Center Children’s Hospital of Pittsburgh. There were no significant differences in demographic characteristics (gender, race, or Hispanic/Latino/Spanish origin) or disease type between participants from the total CCHMC single center and A-LiNK cohort studies and those that met eligibility criteria for this study. In the cohort of eligible participants, 54% were female, with median age at AILD diagnosis of 14 years (range 1 to 23, Table 1). The participants were 86% White and 8% African American/Black. A higher proportion of African American/Black participants were enrolled in non-CCHMC sites (p = 0.03).

**Figure 1:**
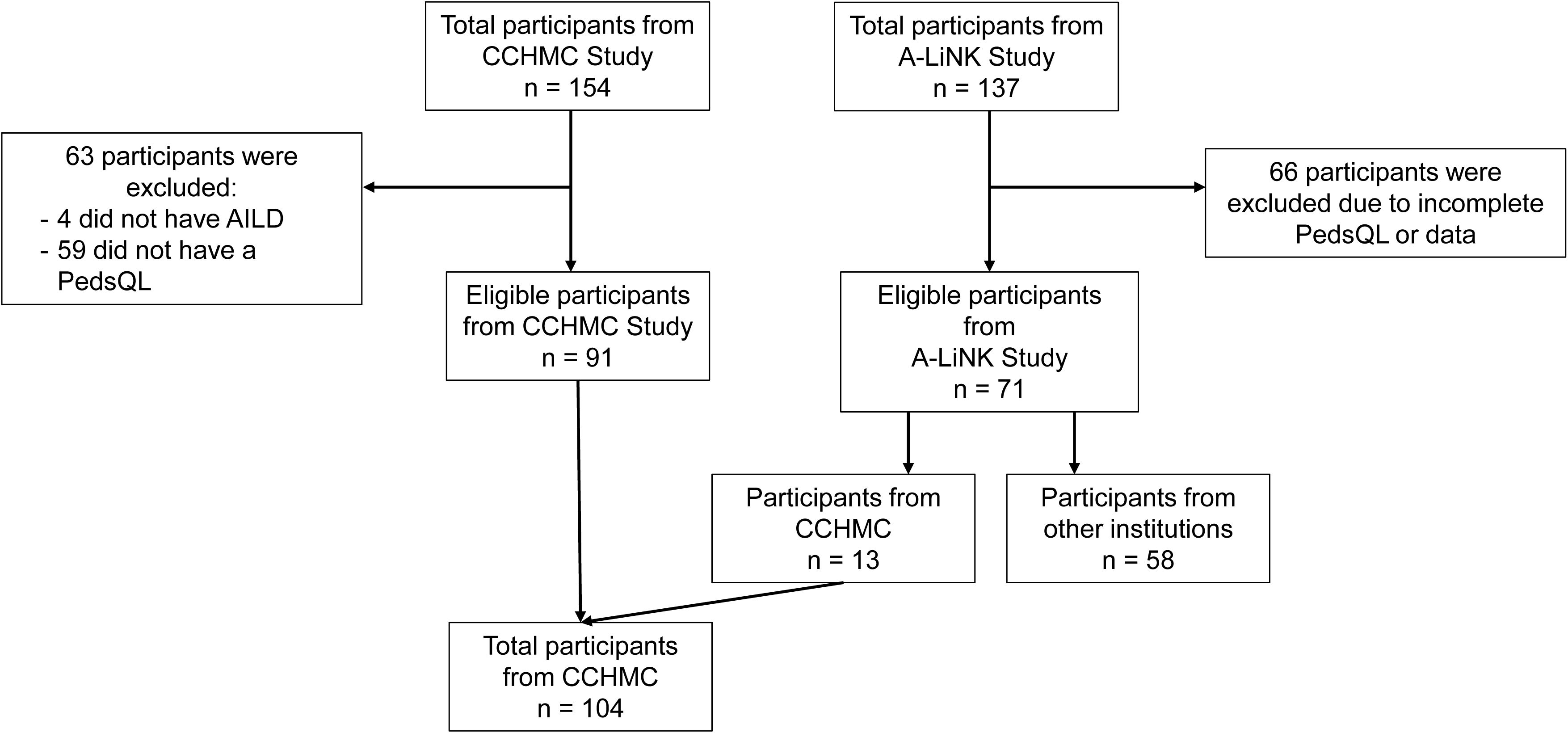
Study CONSORT diagram.

**Table 1:** Demographics, disease characteristics and laboratory values of the cohort at baseline form completion by study site.

|  | All,<br>n = 162 | CCHMC,<br>n = 104 | Other<br>Institutions,<br>n = 58 | *p*<sup>□</sup> |
| --- | --- | --- | --- | --- |
| *Demographics* |  |  |  |  |
| Female gender | 88 (54%) | 53 (51%) | 35 (60%) | 0.25 |
| Age at diagnosis, y | 14 (1, 23) | 14 (1, 23) | 14 (1, 20) | 0.74 |
| Race |  |  |  | **0.03** |
| White | 139 (86%) | 94 (90%) | 45 (78%) |  |
| African American/Black | 13 (8%) | 4 (4%) | 9 (15%) |  |
| Other | 10 (6%) | 6 (6%) | 4 (7%) |  |
| Hispanic/Latino/Spanish<br>origin | 7 (4%) | 5 (5%) | 2 (3%) | 0.13 |
| *Disease Duration, y* | 2 (0, 14) | 2 (0, 14) | 3 (0, 14) | 0.47 |
| *Disease Type* |  |  |  |  |
| AIH | 100 (62%) | 53 (51%) | 47 (81%) | **0.0006** |
| AIH/PSC Overlap | 31 (19%) | 24 (23%) | 7 (12%) |  |
| PSC | 31 (19%) | 27 (26%) | 4 (7%) |  |
| *Comorbidities* |  |  |  |  |
| Inflammatory Bowel<br>Disease | 51 (31%) | 38 (37%) | 13 (22%) | 0.06 |
| Participants with<br>complication(s)<sup>°</sup> | 18 (11%) | 14 (13%) | 4 (7%) | 0.20 |
| *Medications* |  |  |  |  |
| Azathioprine use | 59 (36%) | 38 (37%) | 21 (36%) | 1.0 |
| Median dose<br>(mg/kg/day) | 1.41 (0.49,<br>2.83) | 1.44 (0.49,<br>2.83) | 1.40 (0.58, 2.31) | 0.89 |
| Prednisone use | 55 (34%) | 34 (33%) | 21 (36%) | 0.65 |
| Median dose<br>(mg/kg/day) | 0.16 (0.02,<br>1.23) | 0.16 (0.02,<br>1.23) | 0.16 (0.04, 0.92) | 0.92 |

| ***Labs*** |  |  |  |  |
| --- | --- | --- | --- | --- |
| ALT (U/L) | 39 (6, 3007)<br>n = 149 | 38 (10, 3007)<br>n = 103 | 40 (6, 727)<br>n = 46 | 1.0 |
| AST (U/L) | 33 (10, 2866)<br>n = 149 | 33 (10, 2866)<br>n = 103 | 33 (11, 399)<br>n = 46 | 0.49 |
| Alk Phos (U/L) | 141 (29, 993)<br>n = 149 | 156 (40, 993)<br>n = 103 | 129 (29, 284)<br>n = 46 | **0.03** |
| GGT (U/L) | 30 (4, 594)<br>n = 143 | 34 (6, 521)<br>n = 99 | 30 (4, 594)<br>n = 44 | 0.98 |
| Total bilirubin (mg/dL) | 0.6 (0.1, 33.9)<br>n = 149 | 0.5 (0.1, 33.9)<br>n = 103 | 0.7 (0.1, 2.4)<br>n = 46 | 0.49 |
| Total protein (gm/dL) | 7.5 (5.8, 9.7)<br>n = 148 | 7.5 (5.8, 9.7)<br>n = 103 | 7.4 (6.3, 8.9)<br>n = 45 | 0.91 |
| IgG (mg/dL) | 1320 (618, 3340)<br>n = 70 | 1310 (618, 3340)<br>n = 61 | 1337 (719, 3065)<br>n = 9 | 0.83 |
| Platelets (x10<sup>3</sup>/mcL) | 242 (40, 609)<br>n = 141 | 241 (40, 609)<br>n = 101 | 259 (45, 495)<br>n = 40 | 0.85 |
| INR | 1.1 (0.9, 1.7)<br>n = 77 | 1.1 (0.9, 1.7)<br>n = 64 | 1.1 (0.9, 1.4)<br>n = 13 | 0.16 |
| APRI (points) | 0.35 (0.08, 40.9)<br>n = 141 | 0.36 (0.08, 40.9)<br>n = 101 | 0.35 (0.09, 5.8)<br>n = 40 | 0.84 |
| FIB-4 (points) | 0.37 (0.07, 8.8)<br>n = 141 | 0.36 (0.09, 8.8)<br>n = 101 | 0.43 (0.07, 2.8)<br>n = 40 | 0.28 |
| ***Disease Control*** |  |  |  |  |
| Biochemical Remission | 56 (49%)<br>n = 149 | 49 (48%)<br>n = 103 | 24 (52%)<br>n = 46 | 0.85 |
| GGT < 50 U/L | 82 (57%)<br>n = 143 | 57 (58%)<br>n = 99 | 25 (57%)<br>n = 44 | 0.93 |

Data presented as n (%) or as median (min, max).
°Complications: ascites, cholangitis, esophageal variceal bleeding, hepatic encephalopathy
□ Comparison between CCHMC and other institutions examined using Chi-square/Fisher's exact test or Wilcoxon rank-sum test as applicable.

At the time of the baseline HRQoL form, the median age was 16 years (range 4 to 24), and the median disease duration was 2 years (range 0 to 14, Table 1). The majority of participants had AIH (62%), with the remainder having an equal distribution between PSC (19%) and AIH/PSC overlap syndrome (19%). The latter participants were primarily enrolled at CCHMC. In terms of comorbidities, 31% participants, especially those with biliary tract involvement, had concurrent inflammatory bowel disease, and 11% had an AILD complication. Azathioprine was prescribed to 36% of participants at a median dose of 1.41 mg/kg/day (75 mg/day), and prednisone to 34% of participants at a median dose of 0.16 mg/kg/day (10 mg/day). Median alkaline phosphatase levels were higher at CCHMC compared to non-CCHMC sites. A greater proportion of participants at CCHMC had PSC or overlap syndrome. Participants’ median liver biochemistries were in the upper limits of normal or only mildly elevated. The calculated median APRI and FIB-4 scores were below thresholds commonly associated with advanced fibrosis in AILD, as cut-off values of 0.94 for APRI and 0.47 for FIB-4 have been shown to differentiate mild fibrosis (Metavir F0–F1) from more advanced fibrosis (Metavir F2–F4) in AILD ^18^. In terms of disease control, 49% of participants were in biochemical remission (defined as ALT/AST within the upper limit of normal for age and sex), and 57% had GGT levels less than 50 U/L.

All 162 participants and 139 caregivers completed a baseline PedsQL 4.0 Generic Core Short Form (Table 2). At baseline, compared to healthy controls, participants with AILD reported the lowest mean HRQoL scores in school (73 vs. 79) and emotional (79 vs. 81) domains, while physical (84 vs. 84) and social (89 vs. 87) domain scores were similar to healthy peers. Total and psychosocial scores were also lower than those of healthy controls (81 vs. 83 and 80 vs. 82, respectively) ^14^. As seen in other pediatric chronic diseases, the mean score in the school domain was notably lower than in other domains ^19^. Baseline HRQoL scores were similar between participants from CCHMC and other institutions. Additionally, baseline participant and caregiver scores were moderately correlated, with the highest correlation for physical and total scores (r > 0.6) and lowest correlation for social scores (r = 0.38, Figure 2).

**Figure 2:**
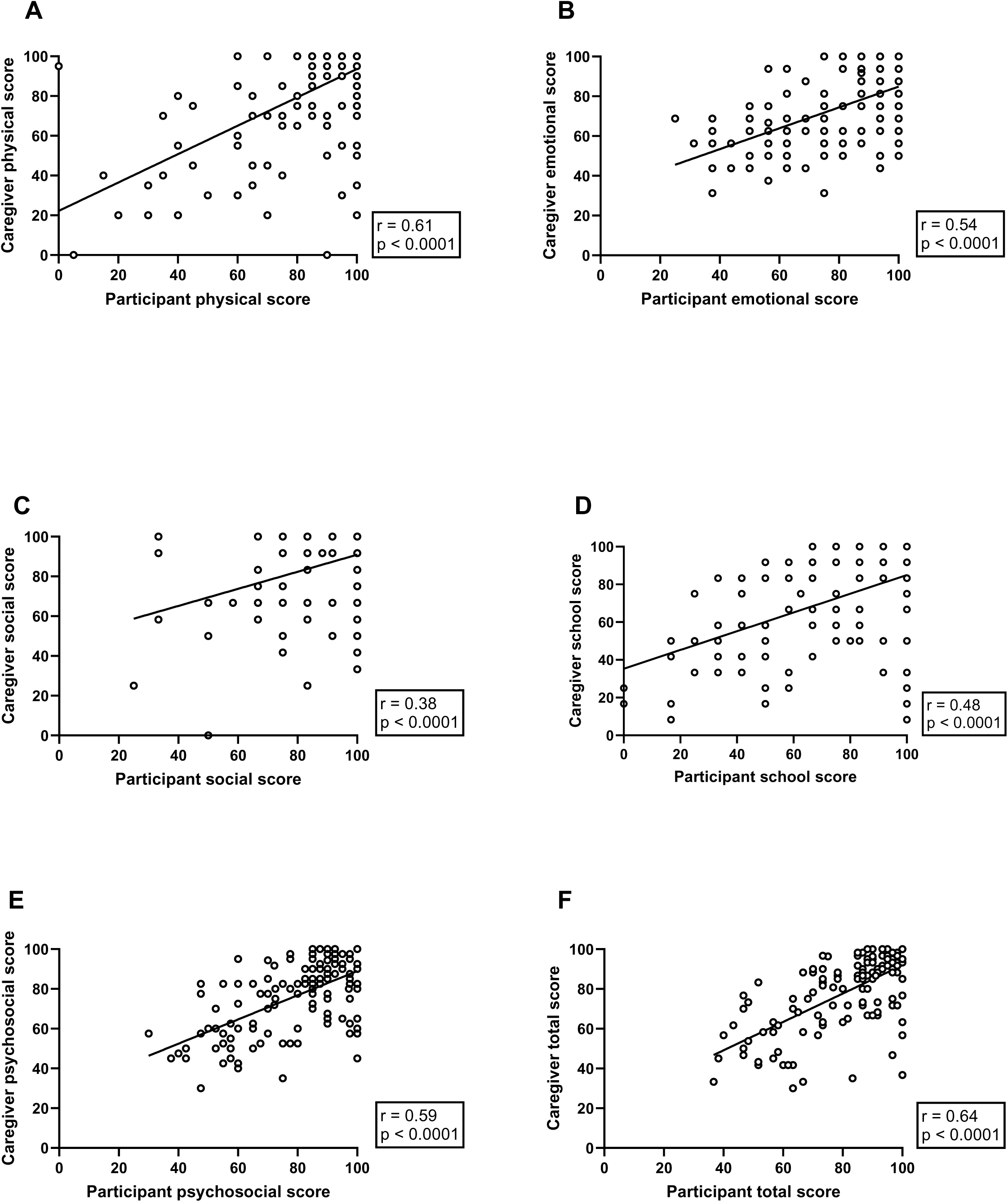
Scatter plots illustrating the relationship between participant and caregiver baseline HRQoL scores. Correlation of participant and caregiver scores from the following domains: (A) physical, (B) emotional, (C) social, (D) school, (E) psychosocial, and (F) total. Pearson correlation coefficients (r) are noted for each scatter plot in the boxes to the right. The black line in each scatter plot represents the best-fit linear regression.

**Table 2:** Baseline HRQoL scores for participants and caregivers.

| *Domain* | *Participant* |  |  |  | *Caregiver* |  |  |  |
| --- | --- | --- | --- | --- | --- | --- | --- | --- |
| | **All,**<br>**n = 162** | **CCHMC,**<br>**n = 104** | **Other**<br>**Institutions,**<br>**n = 58** | *p* □ | **All,**<br>**n = 139** | **CCHMC,**<br>**n = 89** | **Other**<br>**Institutions,**<br>**n = 50** | *p* □ |
| **Physical** | 90 (0, 100)<br>84 ± 22 | 90 (0, 100)<br>82 ± 24 | 95 (15, 100)<br>86 ± 19 | 0.48 | 100 (0, 100)<br>82 ± 26 | 100 (0, 100)<br>81 ± 27 | 100 (0, 100)<br>84 ± 25 | 0.62 |
| **Emotional** | 81 (25, 100)<br>79 ± 20 | 81 (25, 100)<br>78 ± 20 | 81 (31, 100)<br>79 ± 20 | 0.88 | 75 (31, 100)<br>74 ± 19 | 69 (31, 100)<br>72 ± 19 | 75 (31, 100)<br>76 ± 20 | 0.31 |
| **Social** | 100 (25, 100)<br>89 ± 17 | 100 (25, 100)<br>88 ± 18 | 100 (33, 100)<br>92 ± 13 | 0.57 | 100 (0, 100)<br>86 ± 20 | 100 (0, 100)<br>84 ± 22 | 100 (58, 100)<br>90 ± 14 | 0.30 |
| **School** | 75 (0, 100)<br>73 ± 25 | 75 (0, 100)<br>72 ± 26 | 83 (17, 100)<br>74 ± 26 | 0.48 | 75 (8, 100)<br>71 ± 27 | 75 (8, 100)<br>72 ± 27 | 75 (8, 100)<br>71 ± 27 | 0.72 |
| **Psychosocial** | 85 (30, 100)<br>80 ± 17 | 85 (30, 100)<br>79 ± 18 | 85 (40, 100)<br>82 ± 17 | 0.42 | 83 (30, 100)<br>77 ± 18 | 80 (30, 100)<br>76 ± 19 | 83 (40, 100)<br>79 ± 16 | 0.41 |
| **Total** | 87 (37, 100)<br>81 ± 16 | 87 (37, 100)<br>80 ± 17 | 87 (38, 100)<br>83 ± 16 | 0.27 | 85 (30, 100)<br>79 ± 19 | 85 (30, 100)<br>77 ± 20 | 85 (35, 100)<br>80 ± 18 | 0.48 |

*Data presented as median (min, max) and mean ± standard deviation.*
□ *Comparison between CCHMC and other institutions examined using Wilcoxon rank-sum test.*

Of the 162 participants, 85 completed a second PedsQL form, and 43 completed a third. Among the 139 caregivers, 71 and 36 completed a second and third form, respectively. There were no significant differences in the participant demographics, disease types or co-morbidities across the three time points (Supplemental Table 2). The disease duration was significantly longer at the third time point. By the third form, fewer participants were taking prednisone, ALT and AST levels were lower, and a greater proportion had controlled disease. Trends in HRQoL scores remained consistent across the second and third forms, with the school and emotional domains continuing to be the most impacted. No statistically significant differences were observed in domain or total scores across the three time points (Supplemental Table 3).

Simple longitudinal linear mixed-effect regression models revealed statistically and clinically significant associations between either child or caregiver-reported HRQoL scores and presence of AILD complications, use of azathioprine, use of prednisone, and type of AILD (Figure 3). The presence of AILD complications was associated with higher child-reported school scores. Regarding medications, azathioprine use was associated with lower child-reported social HRQoL scores, whereas prednisone use was linked to higher emotional scores in both child- and caregiver-reported data. When prednisone dosing was categorized based on the median value of 0.16 mg/kg/day (high dose: ≥0.16 mg/kg/day, low dose: <0.16 mg/kg/day, and no use), child-reported emotional scores were significantly higher in the low-dose group (LSMeans 88, 95% CI: 83-94) compared to both the high-dose group (LSMeans 77, 95% CI: 70-84; p = 0.01) and those not receiving prednisone (LSMeans 78, 95% CI: 75-81; p = 0.003). Similarly, caregiver-reported emotional scores were significantly higher with low-dose prednisone (LSMeans 85, 95% CI: 78-91) compared to no use (LSMeans 74, 95% CI: 70-77; p = 0.004). No significant difference was observed between low- and high-dose groups in caregiver scores (LSMeans 75, 95% CI: 68-83; p = 0.10).

**Figure 3:**
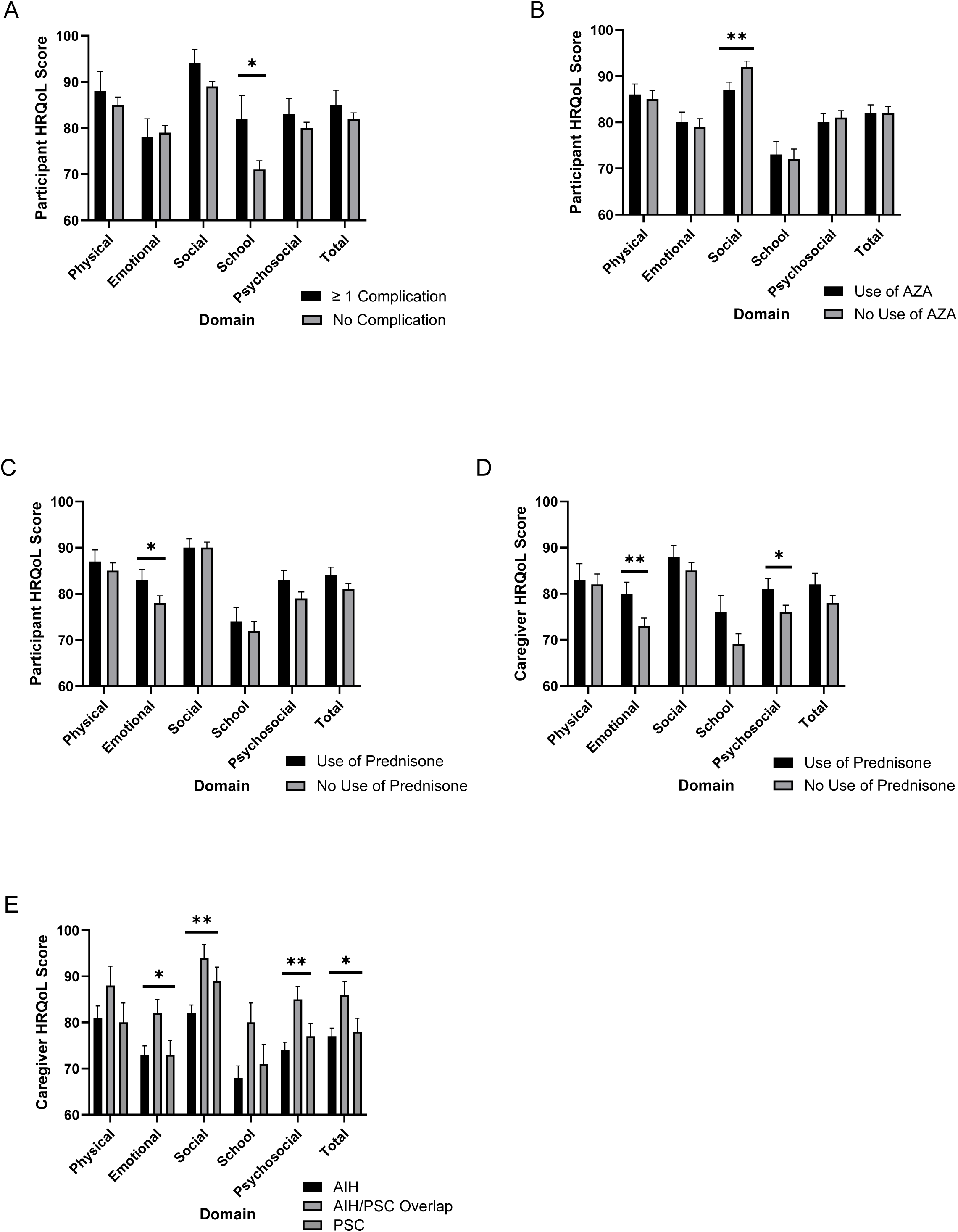
Longitudinal association between HRQoL scores and clinical factors. Association between participant scores and: (A) presence of complications (complication: 18 participants, 34 observations; no complication: 144 participants, 258 observations), (B) use of azathioprine (use: 58 participants, 110 observations; no use: 103 participants, 182 observations), and (C) use of prednisone (use: 55 participants, 79 observations; no use: 107 participants, 213 observations). Association between caregivers scores and: (D) use of prednisone (use: 46 caregivers, 65 observations; no use: 93 caregivers, 181 observations), and (E) type of AILD (AIH: 84 caregivers, 139 observations; Overlap: 28 caregivers, 52 observations; PSC: 28 caregivers, 56 observations). Comparison of HRQoL scores between categories examined longitudinally using linear mixed modeling. Data presented as least squares means ± standard error (indicated by error bars). Significance is denoted with use of asterisk (* = p ≤ 0.05, ** = p ≤ 0.01).

Additionally, from the longitudinal modeling, caregiver-report of HRQoL varied by AILD type, with those caring for participants diagnosed with AIH/PSC overlap syndrome reporting higher emotional, social, psychosocial, and total scores than caregivers of participants with AIH or PSC. All these findings were clinically meaningful, with a minimal clinically important difference of at least five points between scores. There was no significant association between quality of life scores – either total or domain-specific – for participants or caregivers in relation to disease duration or the presence of IBD.

Lastly, simple longitudinal linear mixed-effect regression models revealed significant associations between HRQoL scores and ALT, AST, and GGT levels, while no association was observed with alkaline phosphatase (Figure 4A). For participants, elevated ALT was associated with lower emotional scores, while elevated GGT was associated with lower social scores. For caregivers, elevated ALT was associated with lower scores in five out of six domains: physical, social, school, psychosocial, and total. Additionally, for caregivers, elevated AST was associated with lower social domain scores, and elevated GGT was associated with lower physical and total domain scores. An illustrative graph based on the linear regression model demonstrates that increasing serum ALT levels are associated with domain-specific decreases in participant- or caregiver-reported HRQoL scores (Figure 4B).

**Figure 4:**
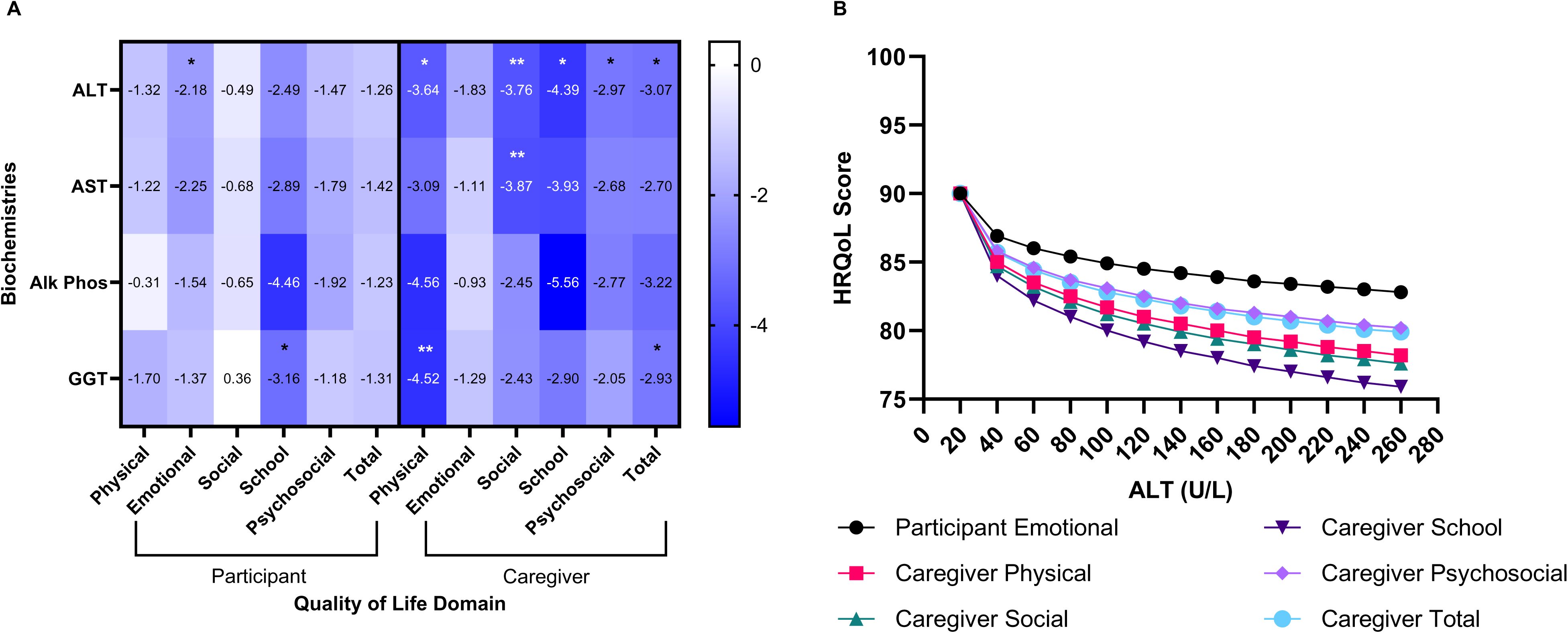
Longitudinal associations between HRQoL scores and biochemical measures. (A) Heat map illustrating the association between HRQoL scores and biochemistries. Values in each cell represent β-estimates; negative β-values indicate that higher biochemistries are associated with lower HRQoL scores. Statistical significance is denoted by asterisks (* = p ≤ 0.05, ** = p ≤ 0.01). (B) Illustrative graph showing the decline in participant- or caregiver-reported HRQoL domain scores with increasing ALT, assuming a baseline quality of life score of 90 at an ALT of 20 U/L.

From the 91 participants enrolled in the CCHMC single center cohort, fatigue was assessed at the time of the baseline form in 70 (77%) participants by either a research survey (n = 61) or from clinic notes (n = 9). Fatigue was present in the majority, 41 (59%) of participants. Pruritus was assessed from clinic notes in 68 (75%) participants and was present in 20 (29%). To assess whether the method of fatigue data collection (survey vs. clinic notes) modified the relationship between fatigue and HRQoL scores, we examined each PedsQL domain longitudinally using models that included fatigue status, collection method, and their interaction. There were no statistically significant interactions, indicating that the association between fatigue and HRQoL scores did not differ by method of data collection.

A significant association was found between age group (<5 years, 5–12 years, >12 years) and presence of fatigue, with fatigue more commonly reported in participants older than 12 years (Supplemental Table 4).

No significant differences in disease characteristics were identified between participants based on presence or absence of fatigue or pruritus (Table 3). However, participants reporting fatigue were diagnosed at an older median age, had significantly higher ALT and AST levels, and were less likely to be in biochemical remission compared to those without fatigue.

**Table 3:** Demographics and disease characteristics of participants from CCHMC single center segregated by presence and absence of symptoms.

|  | Presence<br>of Fatigue<br>n = 41 | Absence of<br>Fatigue<br>n = 29 | *p* <sup>□</sup> | Presence<br>of<br>Pruritus<br>n = 20 | Absence<br>of<br>Pruritus<br>n = 48 | *p* <sup>□</sup> |
| --- | --- | --- | --- | --- | --- | --- |
| *Demographics* |  |  |  |  |  |  |
| Female gender | 23 (56%) | 14 (48%) | 0.52 | 12 (60%) | 23 (48%) | 0.36 |
| Age at diagnosis, y | 14 (1, 23) | 12 (1, 19) | **0.05** | 15 (1, 19) | 13 (2, 23) | 0.36 |
| Race |  |  | 0.46 |  |  | 0.14 |
| White | 36 (88%) | 28 (97%) |  | 16 (80%) | 45 (94%) |  |
| African American/Black | 3 (7%) | 0 (0%) |  | 2 (10%) | 1 (2%) |  |
| Other | 2 (5%) | 1 (3%) |  | 2 (10%) | 2 (4%) |  |
| Hispanic/Latino/Spanish origin | 2 (5%) | 1 (3%) | 1.0 | 2 (10%) | 0 (0%) | 0.08 |
| *Disease Duration, y* | 1 (0, 8) | 2 (0, 8) | 0.52 | 0.5 (0, 8) | 1 (0, 8) | 0.54 |
| *Disease Type* |  |  |  |  |  |  |
| AIH | 22 (54%) | 13 (44%) | 0.69 | 6 (30%) | 24 (50%) | 0.29 |
| AIH/PSC Overlap | 8 (19%) | 8 (28%) |  | 8 (40%) | 12 (25%) |  |
| PSC | 11 (27%) | 8 (28%) |  | 6 (30%) | 12 (25%) |  |
| *Comorbidities* |  |  |  |  |  |  |
| Inflammatory Bowel disease | 13 (32%) | 10 (34%) | 0.81 | 8 (40%) | 18 (38%) | 0.85 |
| Participants with complication(s)<sup>°</sup> | 7 (17%) | 3 (10%) | 0.51 | 5 (25%) | 4 (8%) | 0.11 |
| *Medications* |  |  |  |  |  |  |
| Azathioprine use | 11 (27%) | 12 (41%) | 0.20 | 3 (15%) | 18 (38%) | 0.07 |
| Prednisone use | 12 (29%) | 11 (38%) | 0.45 | 6 (30%) | 17 (35%) | 0.67 |
| *Labs* |  |  |  |  |  |  |
| ALT (U/L) | 66 (10, 2298) | 31 (10, 3007) | **0.02** | 34 (10, 709) | 39 (10, 3007) | 0.83 |
|  | n = 40 | n = 29 |  | n = 20 | n = 47 |  |
| AST (U/L) | 46 (14, 1897)<br>n = 40 | 30 (10, 2866)<br>n = 29 | **0.04** | 39 (14, 583)<br>n = 20 | 32 (10, 2866)<br>n = 47 | 0.99 |
| Alk Phos (U/L) | 141 (45, 433)<br>n = 31 | 177 (50, 575)<br>n = 25 | 0.63 | 195 (45, 433)<br>n = 19 | 142 (50, 575)<br>n = 36 | 0.17 |
| GGT (U/L) | 51 (10, 500)<br>n = 40 | 20 (6, 494)<br>n = 27 | 0.52 | 58 (10, 494)<br>n = 20 | 30 (6, 500)<br>n = 46 | 0.19 |
| Total bilirubin (mg/dL) | 0.6 (0.1, 25.1)<br>n = 40 | 0.4 (0.1, 33.9)<br>n = 29 | 0.17 | 0.5 (0.1, 25.1)<br>n = 20 | 0.5 (0.1, 33.9)<br>n = 47 | 0.89 |
| Platelets (x10<sup>3</sup>/mcL) | 229 (40, 609)<br>n = 39 | 286 (44, 443)<br>n = 28 | 0.21 | 286 (63, 496)<br>n = 20 | 269 (40, 609)<br>n = 45 | 0.55 |
| ***Disease Control*** |  |  |  |  |  |  |
| Biochemical Remission | 13 (33%)<br>n = 40 | 16 (55%)<br>n = 29 | **0.01** | 9 (41%)<br>n = 20 | 17 (36%)<br>n = 47 | 0.5 |
| GGT < 50 | 20 (50%)<br>n = 40 | 18 (67%)<br>n = 27 | 0.18 | 8 (40%)<br>n = 20 | 28 (60%)<br>n = 47 | 0.14 |

*Data presented as n (%) or as median (min, max).*
*°Complications: ascites, cholangitis, esophageal variceal bleeding, hepatic encephalopathy*
*□ Comparison by presence v. absence of symptoms examined using Chi square/Fisher's exact test or Wilcoxon rank-sum test as applicable.*

Fatigue was associated with significantly lower HRQoL scores. Participants with fatigue had reduced physical, emotional, school, psychosocial and total scores (Figure 5). Caregivers of participants with fatigue reported lower scores across all domains. Similarly, participants with pruritus had significantly lower physical, emotional, psychosocial, and total scores, and caregiver-reported total scores were also lower among those with pruritus.

**Figure 5:**
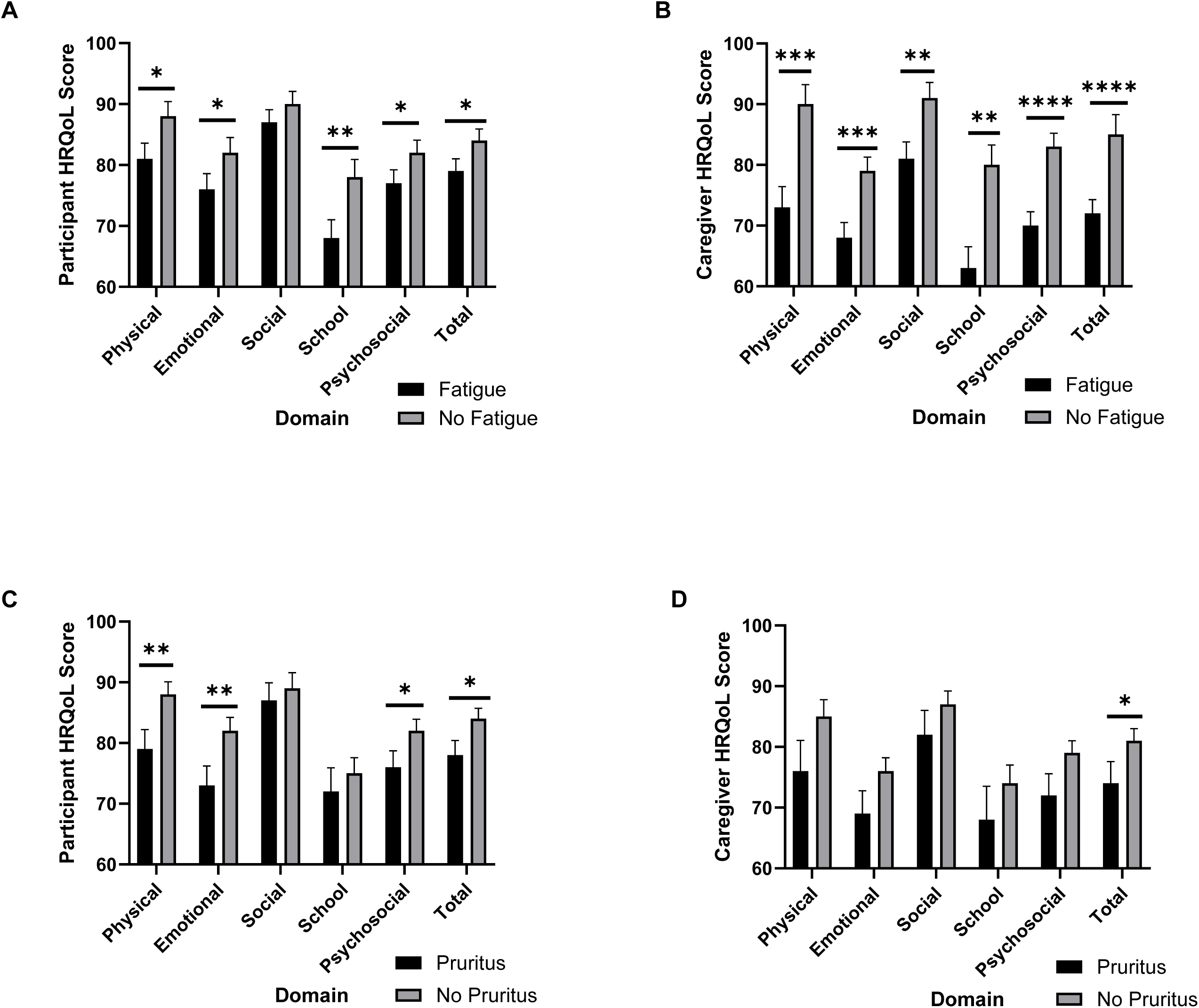
Longitudinal association between HRQoL scores and presence of symptoms. Association between (A) participant scores and fatigue (fatigue: 41 participants, 77 observations; no fatigue: 29 participants, 74 observations) and (B) caregiver scores and fatigue (fatigue: 32 caregivers, 61 observations; no fatigue: 27 caregivers, 70 observations). Association between (C) participant scores and pruritus (pruritus: 20 participants, 34 observations; no pruritus: 48 participants, 115 observations) and (D) caregivers scores and pruritus (pruritus: 15 caregivers, 25 observations; no pruritus: 43 caregivers, 101 observations). Comparison of HRQoL scores between presence v. absence of symptoms examined longitudinally using linear mixed modeling. Data presented as least squares means ± standard error (indicated by error bars). Significance is denoted with the use of asterisk (* = p ≤ 0.05, ** = p ≤ 0.01, *** = p ≤ 0.001, **** = p ≤ 0.0001).

Regarding changes in HRQoL scores over time, there were no significant interactions among most of the clinical factors (presence of AILD complications, presence of IBD, use of azathioprine, or presence of fatigue) and time for any quality of life scores, either total or domain-specific. However, two significant interactions were observed. First, participants taking prednisone had lower baseline school scores (mean = 70), but scores improved in the second and third assessments, both reaching a mean of 85 (p = 0.005); no time-based effect was seen for emotional scores. Second, participants with pruritus had higher baseline emotional scores (mean = 76), but scores declined in the later assessments, dropping to 74 in the second and 58 in the third (p = 0.02).

In multivariable models, AILD complications were associated with higher participant-reported school, psychosocial, and total HRQoL scores (Supplemental Table 5). Among caregiver-proxy reports, caregivers of participants with AIH reported lower social and psychosocial scores compared to those of participants with PSC (Supplemental Table 6). Additionally, prednisone use was associated with higher caregiver-reported psychosocial scores. Fatigue was associated with lower participant-reported school scores and lower caregiver-reported scores across all domains. Pruritus was associated with lower participant-reported physical, emotional, and total scores. Notably, results for fatigue and pruritus from a subset multivariable analysis using data from the CCHMC single-center cohort were consistent with those from the full multivariable models.

## Discussion

This study is the first multicenter, longitudinal study to examine factors associated with quality of life in pediatric AILD and the largest to date in this population. Overall, quality of life was reduced in children with AILD, with the impact varying by domain. Compared to healthy controls, participants had lower mean scores in emotional and school domains, as well as lower psychosocial and total scores. Caregivers reported lower scores in all domains.

The correlation between child and caregiver HRQoL scores in our study was moderate and consistent with previous research, ranging from 0.3 to 0.6. Correlations were lower for subjective domains (e.g., social and emotional functioning) and higher for observable domains (e.g., physical functioning). As reported in other chronic disease populations, caregivers tended to rate children’s HRQoL lower than the children themselves, a discrepancy that may reflect caregiver stress and potentially their own quality of life ^20^. We did not collect data on the caregiver’s relationship with the child (e.g., mother, father, guardian), which may have provided further context. A recent study in pediatric cancer found that while both maternal and paternal proxy reports can offer valid insight into a child’s HRQoL, fathers tend to overestimate HRQoL, particularly in subjective domains, whereas mothers’ reports more closely aligned with the child’s own assessment ^21^.

In the multicenter cohort, simple longitudinal mixed-effects modeling revealed several significant associations between clinical factors and quality of life. Participants taking azathioprine reported lower social scores, though scores remained the same as healthy children, and caregiver reports did not report a significant difference ^20^. It may be, however, participants taking azathioprine have slightly lower social functioning related to adverse drug side effects, such as gastrointestinal upset. Conversely, participants treated with prednisone, as well as their caregivers, reported higher scores in the emotional quality of life domain compared to those not treated with prednisone. When segregated into high and low dosing, participants taking low dose prednisone had the highest emotional scores. Interestingly, a study of patients with rheumatoid arthritis treated with low-dose prednisone at night found that they experienced improved fatigue levels and overall quality of life compared to those receiving a placebo. In this study, nighttime administration of prednisone was timed to align with the rise in nocturnal inflammatory cytokines that contribute to morning stiffness ^22^. Based on these findings, we speculate that prednisone may help control inflammation and its associated symptoms, such as fatigue, in the AILD population, thereby enhancing participants’ quality of life. Alternatively, it is possible that patients not taking prednisone experience greater emotional distress and a reduced quality of life, a phenomenon previously observed in cases of secondary adrenal insufficiency following long-term corticosteroid therapy ^23^. Remarkably, when we evaluated the interaction between prednisone use and time, we found that participant school-related quality of life improved over time, further suggesting a potential benefit of prednisone.

Some of the findings from the multicenter analysis were unexpected, particularly the observation that participants with more complications reported better school functioning. However, it is possible that patients with more complex disease states receive greater support through mechanisms such as increased involvement of social workers or the implementation of individualized education plans, tailored to address their specific needs. Lastly, from the multicenter study, disease duration was not found to be associated with quality of life. This result is not surprising as long-term remission can be challenging to maintain in AIH and there is no definitive cure for PSC ^4,15,24^. There was also no association with the presence of IBD and quality of life.

From the single center cohort study, fatigue and pruritus were strongly associated with impaired quality of life. Gulati et al. similarly found that symptoms contribute to lower overall quality of life scores ^7^. Participants with fatigue, and especially their caregivers, reported lower scores across most domains, with school functioning particularly affected. Fatigue has been recognized as a key driver of decreased quality of life in adult chronic liver disease (CLD) and was recently shown to be a factor in reduced HRQoL in adults with AIH ^12^. The mechanism of fatigue leading to reduced quality of life is postulated to be from systemic inflammation disrupting the liver-brain connection and causing neurotransmitter imbalances in part of the brain that regulates motivation and reward ^25,26^. Consistent with this theory, our study found that elevated ALT values were associated with lower quality of life, particularly in caregiver reports. Additionally, participants with fatigue were more likely to have elevated ALT and AST levels and were less likely to be in biochemical remission. These findings suggest that heightened systemic inflammation may contribute to more profound fatigue. A separate study of adults with CLD also found that fatigue was associated with greater liver inflammation, as indicated by higher median liver biochemistries. AILD was also an independent risk factor for fatigue in this study ^27^. Overall, it is intriguing that caregivers in our study reported a stronger association between elevated ALT levels and reduced quality of life than participants themselves, suggesting that caregivers may be more attuned to the impact of liver inflammation on their child’s overall well-being. Moreover, caregivers consistently reported that fatigue had a greater impact on quality of life than the children did. Fatigue, being a complex and often subjective symptom, may be difficult for children to recognize or articulate, especially in the setting of chronic illness. In contrast, caregivers may be more sensitive to subtle behavioral changes, such as decreased school engagement, that reflect underlying fatigue.

Pruritus, although less prevalent, was also associated with impaired quality of life, particularly in the physical, emotional and psychosocial domains. The proportion of participants reporting pruritus was higher (although not significant) for those with overlap syndrome and PSC, diseases that typically result in more severe cholestasis. In contrast to the pattern observed with fatigue, participants with pruritus report a greater impact on their quality of life than caregivers do. Unlike fatigue, pruritus is a more tangible, physical symptom, and as such, children may be more aware of its immediate discomfort. Additionally, when evaluating the interaction between time and pruritus, we found that participants’ emotional quality of life declined significantly over time, suggesting that the chronic burden of this distressing symptom may negatively impact emotional well-being specifically.

Comparing simple and multivariate analyses, some associations changed. For example, azathioprine use was no longer significant after adjustment, suggesting confounding effects. However, type of AILD, complications, fatigue, and pruritus remained significant, indicating robust independent associations with quality of life.

Our study has several notable strengths. It is the largest study of HRQoL in pediatric AILD and uses the validated PedsQL tool, enabling comparison to healthy controls. The study is also multicentered, thereby allowing us to recruit a diverse participant population that mirrors that of patients affected by pediatric AILD in the United States ^4,28,29^. Additionally, the PedsQL forms from the A-LiNK study were available in multiple languages, including Spanish and Arabic, enhancing accessibility and inclusivity. Despite assessments being collected across multiple centers and from both participants and their caregivers, baseline HRQoL scores did not differ significantly between institutions, and scores were correlated between children and caregivers, reinforcing validity. Finally, one of the most significant strengths of our study is its longitudinal and prospective design. This approach enabled the collection of repeat participant questionnaires over time and facilitated the use of longitudinal mixed-effects modeling. By leveraging this statistical method, we increased statistical power and enhanced our ability to detect effects that might otherwise have gone unnoticed.

Limitations include sample size constraints when stratifying by disease type—most participants had AIH, limiting comparison groups. Fatigue and pruritus data were limited to one center and not assessed with validated scales, restricting symptom quantification. Additionally, data on medication adherence and mental health were not collected, despite their potential influence on disease activity, symptom reporting and overall quality of life, as recently exemplified by the adult study in AIH ^12^. Finally, liver disease activity was inferred from serum biochemistries and complications, as imaging was not collected, limiting assessment of disease severity’s impact on symptoms and quality of life.

In conclusion, this study emphasizes the complexity of managing pediatric AILD, where both disease-related symptoms and treatments all influence quality of life. The observed positive association between prednisone and quality of life raises an important clinical question – should clinicians aim for steroid-free biochemical remission, or might the benefits of low-dose steroids outweigh their known risks? Addressing this uncertainty may require a comparative effectiveness research trial to evaluate outcomes in real-world practice settings. Additionally, the negative association found between fatigue and quality of life highlights the need for subsequent systematic investigation using standardized patient-reported outcome measures, including validated assessments of mental health. A deeper understanding of the pathophysiology of fatigue, potentially linked to liver inflammation and its influence on the liver-brain axis, could allow for the development of targeted intervention, including pharmacotherapies, to alleviate this debilitating symptom. Finally, given the unique disease characteristics and symptoms that shape quality of life in this population, as highlighted in this study, the development of a pediatric AILD-specific assessment of health-related quality of life would be highly beneficial.

## Data Availability

All data produced in the present work are contained in the manuscript

## Acknowledgement

Thank you to A-LiNK Connections, as well as the study participants and research teams at Cincinnati Children’s Hospital Medical Center, Nationwide Children’s Hospital, Seattle Children’s Hospital, St. Louis Children’s Hospital - Washington University, and University of Pittsburgh Medical Center Children’s Hospital of Pittsburgh.

## List of Abbreviations

AIH: autoimmune hepatitis
AILD: autoimmune liver disease
A-LiNK: Autoimmune Liver disease Network for Kids
ALT: alanine aminotransferase
APRI: AST to platelet ratio index
ASC: autoimmune sclerosing cholangitis
AST: aspartate aminotransferase
CCHMC: Cincinnati Children’s Hospital Medical Center
CCTST: Center for Clinical & Translational Science & Training
CLD: chronic liver disease
FIB-4: Fibrosis-4
GGT: gamma-glutamyl transpeptidase
HRQoL: health-related quality of life
IBD: inflammatory bowel disease
IgG: immunoglobulin G
INR: International Normalized Ratio
MCID: minimal clinically important difference
PedsQL 4.0: Pediatric Quality of Life Inventory™ Version 4.0
PSC: primary sclerosing cholangitis
REDCap: Research Electronic Data Capture

## Notes

**Conflicts of Interest:** Conflicts of interest: nothing to report.

### Competing Interest Statement

The authors have declared no competing interest.

### Funding Statement

This study was funded by Center for Autoimmune Liver Disease at Cincinnati Children's, NIH P30 DK078392, R01 DK095001

### Author Declarations

Ethics committee/IRB of Cincinnati Children's Hospital Medical Center gave ethical approval for this work

### Summary of Updates

Introduction updated to add literature of adult autoimmune liver disease. Results updated to include additional analysis. Figure 1, 3, 5 revised. Added supplementary tables.

