## Supplemental Tables for "Longitudinal, Multicenter Study of Clinical Factors Impacting Health-Related Quality of Life in Pediatric Autoimmune Liver Disease"

**Supplemental Table 1: Demographics of included vs. excluded participants by study site.**

|  | **CCHMC Study** | | | **A-LiNK Study** | | |
| --- | --- | --- | --- | --- | --- | --- |
|  | **Included,**  **n = 91** | **Excluded,**  **n = 63** | ***p^ǂ^*** | **Included,**  **n = 71** | **Excluded,**  **n = 66** | ***p^ǂ^*** |
| Female gender | 47 (52%) | 20 (32%) | **0.02** | 41 (58%) | 38 (58%) | 1.0 |
| Race  White  African American/Black  Other | 82 (90%)  4 (4%)  5 (6%) | 50 (80%)  4 (6%)  9 (14%) | 0.14 | 57 (63%)  9 (13%)  5 (7%) | 52 (79%)  6 (9%)  8 (12%) | 0.51 |
| Hispanic/Latino/Spanish origin | 4 (4%) | 2 (3%) | 1.0 | 3 (4%) | 1 (2%) | 0.62 |

*Data presented as n (%).*

*ǂComparison between participants included and excluded examined using Chi-square/Fisher’s exact test.*

**Supplemental Table 2: Demographics, disease characteristics and laboratory values by form number.**

|  | **Form 1,**  **n = 162** | **Form 2,**  **n = 85** | **Form 3,**  **n = 43** | ***p^ǂ^*** |
| --- | --- | --- | --- | --- |
| ***Demographics*** | | | | |
| Female gender | 88 (54%) | 44 (52%) | 23 (53%) | 0.93 |
| Age at diagnosis, y | 14 (1, 23) | 13 (1, 23) | 13 (1, 23) | 0.40 |
| Race  White  African American/Black  Other | 139 (86%)  13 (8%)  10 (6%) | 74 (87%)  4 (5%)  7 (8%) | 38 (88%)  2 (5%)  3 (7%) | 0.81 |
| Hispanic/Latino/Spanish origin | 7 (4%) | 3 (4%) | 0 (0%) | 0.39 |
| ***Disease Duration, y*** | 2 (0, 14) | 3 (0, 15) | 4 (1, 16) | **0.0003** |
| ***Disease Type*** | | | | |
| AIH | 100 (62%) | 43 (51%) | 23 (53%) | 0.49 |
| AIH/PSC Overlap | 31 (19%) | 22 (26%) | 9 (21%) |  |
| PSC | 31 (19%) | 20 (23%) | 11 (26%) |  |
| ***Comorbidities*** | | | | |
| Inflammatory Bowel  Disease | 51 (31%) | 35 (41%) | 15 (35%) | 0.32 |
| Participants with  complication(s)*°* | 18 (11%) | 11 (13%) | 5 (12%) | 0.91 |
| ***Medications*** | | | | |
| Azathioprine use | 59 (36%) | 37 (44%) | 14 (33%) | 0.40 |
| Median dose (mg/kg/day) | 1.41 (0.49, 2.83) | 1.27 (0.44, 2.49) | 1.44 (0.57, 2.43) | 0.66 |
| Prednisone use | 55 (34%) | 17 (20%) | 7 (16%) | **0.01** |
| Median dose (mg/kg/day) | 0.16 (0.02, 1.23) | 0.09 (0.03, 1.01) | 0.09 (0.06, 0.70) | 0.13 |
| ***Labs*** | | | | |
| ALT (U/L) | 39 (6, 3007)  n = 149 | 28 (7, 447)  n = 83 | 29 (8, 173)  n = 42 | **0.003** |
| AST (U/L) | 33 (10, 2866)  n = 149 | 26 (12, 168)  n = 83 | 27 (15, 300)  n = 42 | **0.02** |
| Alk Phos (U/L) | 141 (29, 993)  n = 149 | 133 (31, 539)  n = 83 | 129 (34, 727)  n = 42 | 0.46 |
| GGT (U/L) | 30 (4, 594)  n = 143 | 28 (3, 206)  n = 80 | 23 (5, 461)  n = 37 | 0.08 |
| Total bilirubin (mg/dL) | 0.6 (0.1, 33.9)  n = 149 | 0.5 (0.1, 4.3)  n = 81 | 0.5 (0.2, 3.1)  n = 41 | 0.72 |
| Total protein (gm/dL) | 7.5 (5.8, 9.7)  n = 148 | 7.4 (5.3, 10.2)  n = 80 | 7.4 (5.7, 9.4)  n = 41 | 0.23 |
| IgG (mg/dL) | 1320 (618, 3340)  n = 70 | 1220 (566, 2620)  n = 38 | 11190 (644, 2882)  n = 20 | 0.36 |
| Platelets (x10^3^/mcL) | 242 (40, 609)  n = 141 | 263 (14, 309)  n = 82 | 261 (35, 433)  n = 39 | 0.89 |
| INR | 1.1 (0.9, 1.7)  n = 77 | 1.1 (0.9, 1.5)  n = 36 | 1.1 (0.9, 1.4)  n = 21 | 0.46 |
| APRI (points) | 0.35 (0.08, 40.9)  n = 141 | 0.30 (0.09, 4.8)  n = 82 | 0.27 (0.10, 4.0)  n = 39 | 0.07 |
| FIB-4 (points) | 0.37 (0.07, 8.8)  n = 141 | 0.35 (1.0, 7.9)  n = 82 | 0.40 (0.12, 3.4)  n = 39 | 0.81 |
| ***Disease Control*** | | | | |
| Biochemical Remission | 56 (49%)  n = 149 | 53 (64%)  n = 83 | 27 (64%)  n = 42 | **0.0001** |
| GGT < 50 U/L | 82 (57%)  n = 143 | 63 (79%)  n = 80 | 28 (76%)  n = 37 | **0.002** |

*Data presented as n (%) or as median (min, max).*

*°Complications: ascites, cholangitis, esophageal variceal bleeding, hepatic encephalopathy*

*ǂComparison between forms examined using Chi-square/Fisher’s exact test or Wilcoxon rank-sum test as applicable.*

**Supplemental Table 3: HRQoL scores for participants and caregivers by form number.**

*Data presented as median (min, max) and mean ± standard deviation.*

*ǂComparison between forms examined using Wilcoxon rank-sum test.*

| ***Domain*** | ***Participant*** | | | | ***Caregiver*** | | | |
| --- | --- | --- | --- | --- | --- | --- | --- | --- |
|  | **Form 1,**  **n = 162** | **Form 2,**  **n = 85** | **Form 3,**  **n = 43** | ***p****ǂ* | **Form 1,**  **n = 139** | **Form 2,**  **n = 71** | **Form 3,**  **n = 36** | ***p****ǂ* |
| **Physical** | 90 (0, 100)  84 ± 22 | 95 (10, 100)  84 ± 22 | 95 (45, 100)  87 ± 16 | 0.69 | 100 (0, 100)  82 ± 26 | 95 (5, 100)  78 ± 29 | 100 (10, 100)  86 ± 25 | 0.36 |
| **Emotional** | 81 (25, 100)  79 ± 20 | 81 (25, 100)  80 ± 20 | 75 (31, 100)  79 ± 19 | 0.88 | 75 (31, 100)  74 ± 19 | 75 (6, 100)  74 ± 20 | 75 (25, 100)  75 ± 19 | 0.88 |
| **Social** | 100 (25, 100)  89 ± 17 | 100 (50, 100)  91 ± 13 | 92 (42, 100)  87 ± 17 | 0.27 | 100 (0, 100)  86 ± 20 | 92 (25, 100)  85 ± 20 | 92 (42, 100)  85 ± 18 | 0.64 |
| **School** | 75 (0, 100)  73 ± 25 | 83 (0, 100)  75 ± 24 | 75 (8, 100)  70 ± 25 | 0.57 | 75 (8, 100)  71 ± 27 | 75 (0, 100)  71 ± 28 | 75 (0, 100)  71 ± 30 | 0.90 |
| **Psychosocial** | 85 (30, 100)  80 ± 17 | 83 (25, 100)  81 ± 17 | 78 (38, 100)  78 ± 16 | 0.62 | 83 (30, 100)  77 ± 18 | 78 (25, 100)  76 ± 18 | 79 (35, 100)  77 ± 18 | 0.95 |
| **Total** | 87 (37, 100)  81 ± 16 | 85 (23, 100)  82 ± 15 | 83 (48, 100)  82 ± 14 | 0.84 | 85 (30, 100)  79 ± 19 | 80 (35, 100)  77 ± 19 | 83 (30, 100)  80 ± 17 | 0.69 |

**Supplemental Table 4:** Number and percentage of participants in each age group (<5 years, 5–12 years, >12 years) stratified by report of fatigue (Yes v. No).

|  | **Yes**  **n = 41** | **No**  **n = 29** | ***pǂ*** |
| --- | --- | --- | --- |
| <5 years | 0 (0%) | 1 (3%) | **0.007** |
| 5-12 years | 5 (12%) | 12 (42%) |  |
| >12 years | 36 (88%) | 16 (55%) |  |

*Data presented as n (%).*

*ǂComparison between age groups examined using Chi-square/Fisher’s exact test.*

**Supplemental Table 5: Comparison of Clinical Factor Associations with Participant HRQoL Scores in Simple and Multivariable Models.**

|  | | **Simple Model** | | | | **Multivariable Model** | | | |
| --- | --- | --- | --- | --- | --- | --- | --- | --- | --- |
| **Clinical Factor** | **HRQoL Domain** | **β-estimate (95% CI)** | **LSMeans** | **LSMeans Diff^1^** | ***p*** | **β-estimate (95% CI)** | **LSMeans** | **LSMeans Diff^1^** | ***p*** |
| **Disease Duration** | Physical | 0.44 (−0.55, 1.43) | — | — | 0.38 | 0.40 (−1.04, 1.84) | — | — | 0.58 |
|  | Emotional | −0.26 (−1.18, 0.67) | — | — | 0.59 | −0.08 (−1.61, 1.44) | — | — | 0.91 |
|  | Social | −0.23 (−0.93, 0.47) | — | — | 0.52 | 0.04 (−1.20, 1.28) | — | — | 0.95 |
|  | School | −0.22 (−1.37, 0.94) | — | — | 0.71 | −0.59 (−2.38, 1.19) | — | — | 0.51 |
|  | Psychosocial | −0.17 (−0.95, 0.62) | — | — | 0.68 | 0.13 (−1.15, 1.40) | — | — | 0.84 |
|  | Total | 0.05 (−0.69, 0.80) | — | — | 0.89 | 0.10 (−1.06, 1.25) | — | — | 0.87 |
| **Type of AILD (reference: PSC)** | Physical  AIH  Overlap  PSC | 1.80 (−5.77, 9.38)  8.16 (−0.58, 16.9)  Ref | 84 vs. 82  91 vs. 82  82 | +2  +9 | 0.13 | −11.95 (−24.06, 0.17)  −4.74 (−15.83, 6.34) Ref | 81 vs. 93  88 vs. 93  93 | −12  −5 | 0.13 |
|  | Emotional  AIH  Overlap  PSC | 3.90 (3.23, 11.04)  6.95 (−1.33, 15.23)  Ref | 79 vs. 76  82 vs. 76  76 | +3  +6 | 0.25 | 9.26 (−3.56, 22.07)  8.83 (−2.97, 20.62)  Ref | 82 vs. 72  81 vs. 72  72 | +10  +9 | 0.27 |
|  | Social  AIH  Overlap  PSC | −1.41 (−6.80, 3.97)  1.16 (−5.23, 7.54) Ref | 89 vs. 90  92 vs. 90  90 | −1  +2 | 0.61 | −4.14 (−14.58, 6.30)  −1.71 (−11.28, 7.86)  Ref | 85 vs. 89  88 vs. 89  89 | −4  −1 | 0.71 |
|  | School  AIH  Overlap  PSC | 6.51 (−2.39, 15.40)  9.03 (−1.32, 19.38)  Ref | 73 vs. 67  76 vs. 67  67 | +6  +9 | 0.20 | 6.96 (−8.07, 21.99)  2.72 (−11.10, 16.54)  Ref | 84 vs. 77  80 vs. 77  77 | +7  +3 | 0.63 |
|  | Psychosocial  AIH  Overlap  PSC | 3.27 (−2.78, 9.32)  5.82 (−1.16, 12.81)  Ref | 80 vs. 77  83 vs. 77  77 | +3  +6 | 0.26 | 4.27 (−6.43, 14.97)  3.89 (−5.95, 13.73)  Ref | 83 vs. 79  83 vs. 79  79 | +4  +4 | 0.68 |
|  | Total  AIH  Overlap  PSC | 3.34 (−2.39, 9.06)  6.98 (0.43, 13.53)  Ref | 82 vs. 79  86 vs. 79  79 | +3  +7 | 0.11 | −0.83 (−10.52, 8.86)  1.23 (−7.68, 10.15)  Ref | 83 vs. 84  85 vs. 84  84 | −1  +1 | 0.87 |
| **AILD Complications**  **(Yes vs. No)** | Physical | 3.22 (−5.69, 12.12) | 88 vs. 85 | +3 | 0.48 | 6.32 (−4.85, 17.48) | 90 vs. 84 | +6 | 0.26 |
|  | Emotional | −1.83 (−10.25, 6.58) | 78 vs. 79 | −1 | 0.67 | 4.89 (−6.88, 16.67) | 81 vs. 76 | +5 | 0.41 |
|  | Social | 5.01 (−1.34, 11.37) | 94 vs. 89 | +5 | 0.12 | 5.32 (−4.28, 14.92) | 90 vs. 85 | +5 | 0.27 |
|  | School | 10.22 (−0.16, 20.59) | 82 vs. 71 | +11 | **0.05** | 22.22 (8.43, 36.01) | 91 vs. 69 | +22 | **0.002** |
|  | Psychosocial | 3.45 (−3.64, 10.53) | 83 vs. 80 | +3 | 0.34 | 11.00 (1.14, 20.86) | 87 vs. 76 | +11 | **0.03** |
|  | Total | 3.14 (−3.54, 9.83) | 85 vs. 82 | +3 | 0.35 | 9.42 (0.51, 18.33) | 89 vs. 79 | +10 | **0.04** |
| **IBD**  **(Yes vs. No)** | Physical | 3.16 (−2.94, 9.27) | 87 vs. 84 | +3 | 0.31 | −6.78 (−16.98, 3.42) | 84 vs. 90 | −6 | 0.19 |
|  | Emotional | 1.35 (−4.41, 7.12) | 80 vs. 79 | +1 | 0.64 | 4.70 (−5.91, 15.32) | 81 vs. 76 | +5 | 0.38 |
|  | Social | 1.45 (−2.94, 5.84) | 91 vs. 89 | +2 | 0.51 | −4.28 (−13.05, 4.50) | 85 vs. 90 | −5 | 0.33 |
|  | School | 0.47 (−6.71, 7.65) | 73 vs. 72 | +1 | 0.90 | −0.98 (−13.50, 11.54) | 80 vs. 81 | −1 | 0.88 |
|  | Psychosocial | 0.69 (−4.18, 5.56) | 81 vs. 80 | +1 | 0.78 | 0.69 (−8.09, 9.47) | 82 vs. 81 | +1 | 0.87 |
|  | Total | 1.24 (−3.35, 5.84) | 83 vs. 82 | +1 | 0.59 | −2.16 (−10.16, 5.83) | 83 vs. 85 | −2 | 0.59 |
| **Prednisone**  **(Yes vs. No)** | Physical | 1.96 (−3.34, 7.25) | 87 vs. 85 | +2 | 0.47 | 4.20 (−4.15, 12.56) | 89 vs. 85 | +4 | 0.32 |
|  | Emotional | 5.28 (0.24, 10.33) | 83 vs. 78 | +5 | **0.04** | 4.13 (−3.72, 11.97) | 80 vs. 76 | +4 | 0.29 |
|  | Social | 0.20 (−4.06, 4.45) | 90 vs. 90 | 0 | 0.93 | −1.66 (−8.74, 5.42) | 87 vs. 88 | −1 | 0.64 |
|  | School | 2.54 (−3.88, 8.97) | 74 vs. 72 | +2 | 0.43 | 4.51 (−4.96, 13.99) | 83 vs. 78 | +5 | 0.34 |
|  | Psychosocial | 3.94 (−0.27, 8.14) | 83 vs. 79 | +4 | 0.07 | 4.53 (−1.76, 10.82) | 84 vs. 79 | +5 | 0.15 |
|  | Total | 3.12 (−0.75, 7.00) | 84 vs. 81 | +3 | 0.11 | 4.11 (−1.72, 9.94) | 86 vs. 82 | +4 | 0.16 |
| **Azathioprine**  **(Yes vs. No)** | Physical | 1.18 (−4.40, 6.76) | 86 vs. 85 | +1 | 0.68 | −2.54 (−11.29, 6.20) | 86 vs. 88 | −2 | 0.56 |
|  | Emotional | 0.87 (−4.43, 6.17) | 80 vs. 79 | +1 | 0.74 | −4.39 (−13.10, 4.33) | 76 vs. 80 | −4 | 0.32 |
|  | Social | −5.25 (−9.35, −1.14) | 87 vs. 92 | −5 | **0.01** | −6.23 (−13.70, 1.25) | 84 vs. 91 | −7 | 0.10 |
|  | School | 1.31 (−5.33, 7.95) | 73 vs. 72 | +1 | 0.70 | −1.37 (−11.76, 9.01) | 80 vs. 81 | −1 | 0.79 |
|  | Psychosocial | −0.93 (−5.38, 3.53) | 80 vs. 81 | −1 | 0.68 | −4.28 (−11.40, 2.83) | 79 vs. 84 | −5 | 0.23 |
|  | Total | 0.07 (−4.10, 4.23) | 82 vs. 82 | 0 | 0.97 | −2.77 (−9.30, 3.76) | 83 vs. 86 | −3 | 0.40 |
| **Fatigue**  **(Yes vs. No)** | Physical | −7.25 (−13.39, −1.10) | 81 vs. 88 | −7 | **0.02** | −6.17 (−13.22, 0.88) | 84 vs. 90 | −6 | 0.08 |
|  | Emotional | −5.81 (-11.69, 0.07) | 76 vs. 82 | −6 | **0.05** | −4.39 (−10.92, 2.15) | 76 vs. 80 | −4 | 0.18 |
|  | Social | −2.61 (−7.86, 2.64) | 87 vs. 90 | −3 | 0.33 | −2.05 (−8.02, 3.92) | 86 vs. 88 | −2 | 0.49 |
|  | School | −9.64 ( −16.8, −2.48) | 68 vs. 78 | −10 | **0.01** | −9.48 (−17.40, −1.55) | 76 vs. 85 | −9 | **0.02** |
|  | Psychosocial | −5.26 (−10.14, −0.37) | 77 vs. 82 | −5 | **0.04** | −4.49 (−9.70, 0.71) | 79 vs. 84 | −5 | 0.09 |
|  | Total | −5.28 (−9.68, −0.88) | 79 vs. 84 | −5 | **0.02** | −3.93 (−8.78, 0.91) | 82 vs. 86 | −4 | 0.11 |
| **Pruritus**  **(Yes vs. No)** | Physical | −9.04 (−16.14, −1.94) | 79 vs. 88 | −9 | **0.01** | −8.75 (−17.10, −0.40) | 83 vs. 91 | −8 | **0.04** |
|  | Emotional | −9.05 (−15.34, −2.77) | 73 vs. 82 | −9 | **0.01** | −9.56 (−17.06, −2.05) | 74 vs. 83 | −9 | **0.01** |
|  | Social | −2.20 (−8.25, 3.84) | 87 vs. 89 | −2 | 0.47 | −2.77 (−9.80, 4.27) | 86 vs. 89 | −3 | 0.43 |
|  | School | −2.49 (−10.58, 5.59) | 72 v. 75 | −3 | 0.54 | −1.09 (−10.25, 8.07) | 80 vs. 81 | −1 | 0.81 |
|  | Psychosocial | −5.54 (−10.89, −0.19) | 76 vs. 82 | −6 | **0.04** | −5.65 (−11.58, 0.28) | 79 vs. 84 | −5 | 0.06 |
|  | Total | −6.00 (−10.85, −1.15) | 78 vs. 84 | −6 | **0.02** | −5.60 (−11.15, −0.06) | 81 vs. 87 | −6 | **0.05** |

^1^LSMean Diff is the difference between group-adjusted means (Yes – No).

**Supplemental Table 6: Comparison of Clinical Factor Associations with Caregiver HRQoL Scores in Simple and Multivariable Models.**

|  | | **Simple Model** | | | | **Multivariable Model** | | | |
| --- | --- | --- | --- | --- | --- | --- | --- | --- | --- |
| **Clinical Factor** | **HRQoL Domain** | **β-estimate (95% CI)** | **LSMeans** | **LSMeans Diff^1^** | ***p*** | **β-estimate (95% CI)** | **LSMeans** | **LSMeans Diff^1^** | ***p*** |
| **Disease Duration** | Physical | 0.79 (−0.57, 2.15) | — | — | 0.25 | −0.58 (−2.63, 1.47) | — | — | 0.57 |
|  | Emotional | 0.19 (−0.82, 1.21) | — | — | 0.71 | −0.24 (−1.68, 1.19) | — | — | 0.73 |
|  | Social | 0.22 (−0.76, 1.21) | — | — | 0.65 | −0.29 (−1.84, 1.27) | — | — | 0.71 |
|  | School | 0.13 (−1.26, 1.51) | — | — | 0.86 | −0.01 (−2.15, 2.13) | — | — | 0.99 |
|  | Psychosocial | 0.18 (−0.75, 1.11) | — | — | 0.70 | −0.17 (−1.49, 1.15) | — | — | 0.79 |
|  | Total | 0.43 (−0.53, 1.4) | — | — | 0.38 | −0.32 (−1.67, 1.04) | — | — | 0.64 |
| **Type of AILD (reference: PSC)** | Physical  AIH  Overlap  PSC | 0.88 (−8.95, 10.7)  8.33 (−3.18, 19.84)  Ref | 81 vs. 80  88 vs. 80  80 | +1  +8 | 0.25 | −6.04 (−24.11, 12.03)  0.47 (−15.27, 16.21) Ref | 79 vs. 85  85 vs. 85  85 | −6  0 | 0.64 |
|  | Emotional  AIH  Overlap  PSC | −0.45 (−7.62, 6.72)  8.62 (0.21, 17.03)  Ref | 73 vs. 73  82 vs. 73  73 | 0  +9 | **0.03** | −8.12 (−20.73, 4.49)  −0.10 (−11.10, 10.89)  Ref | 69 vs. 77  77 vs. 77  77 | −8  0 | 0.25 |
|  | Social  AIH  Overlap  PSC | −6.26 (−13.18, 0.65)  5.79 (−2.34, 13.91)  Ref | 82 vs. 89  94 vs. 89  89 | −7  +5 | **0.002** | −22.35 (−36.00, −8.71)  −4.85 (−16.77, 7.06)  Ref | 77 vs. 99  94 vs. 99  99 | −22  −5 | **0.003** |
|  | School  AIH  Overlap  PSC | −3.55 (−13.45, 6.35)  8.44 (−3.26, 20.14)  Ref | 68 vs. 71  80 vs. 71  71 | −3  +9 | 0.06 | −16.94 (−35.73, 1.84)  −5.15 (−21.54, 11.24)  Ref | 70 vs. 87  82 vs. 87  87 | −17  −5 | 0.16 |
|  | Psychosocial  AIH  Overlap  PSC | −3.03 (−9.53, 3.47)  7.60 (0, 15.20)  Ref | 74 vs. 77  85 vs. 77  77 | −3  +8 | **0.01** | −14.55 (−26.16, −2.95)  −2.88 (−13.01, 7.25)  Ref | 72 vs. 86  83 vs. 86  86 | −14  −3 | **0.02** |
|  | Total  AIH  Overlap  PSC | −1.70 (−8.54, 5.13)  7.53 (−0.40, 15.46)  Ref | 77 vs. 78  86 vs. 78  78 | −1  +8 | **0.02** | −11.72 (−23.61, 0.17)  −1.75 (−12.12, 8.62)  Ref | 74 vs. 86  84 vs. 86  86 | −12  −2 | 0.08 |
| **AILD Complications**  **(Yes vs. No)** | Physical | −1.22 (−12.95, 10.51) | 81 vs. 82 | −2 | 0.84 | −6.03 (−21.47, 9.41) | 80 vs. 86 | −6 | 0.43 |
|  | Emotional | −1.51 (−10.25, 7.22) | 73 vs. 75 | −2 | 0.73 | −1.19 (−11.97, 9.59) | 74 vs. 75 | −1 | 0.82 |
|  | Social | 2.66 (−5.83, 11.16) | 89 vs. 86 | +3 | 0.54 | 6.66 (−5.00, 18.33) | 93 vs. 86 | +7 | 0.25 |
|  | School | 8.69 (−3.27, 20.65) | 79 vs. 70 | +9 | 0.15 | 10.24 (−5.82, 26.29) | 85 vs. 74 | +11 | 0.20 |
|  | Psychosocial | 2.64 (−5.35, 10.62) | 79 vs. 77 | +2 | 0.51 | 4.64 (−5.28, 14.56) | 83 vs. 78 | +5 | 0.35 |
|  | Total | 1.5 (−6.76, 9.77) | 80 vs. 79 | +1 | 0.72 | 1.02 (−9.14, 11.18) | 82 vs. 81 | +1 | 0.84 |
| **IBD**  **(Yes vs. No)** | Physical | 4.46 (−3.65, 12.58) | 85 vs. 81 | +4 | 0.28 | 1.15 (−13.83, 16.14) | 83 vs. 82 | +1 | 0.88 |
|  | Emotional | 2.49 (−3.57, 8.55) | 76 vs. 74 | +2 | 0.42 | −0.36 (−10.81, 10.10) | 74 vs. 74 | 0 | 0.94 |
|  | Social | 4.12 (−1.72, 9.95) | 89 vs. 85 | +4 | 0.16 | −6.81 (−18.12, 4.50) | 86 vs. 93 | −7 | 0.23 |
|  | School | 7.31 (−0.88, 15.51) | 76 vs. 68 | +8 | 0.08 | −2.94 (−18.52, 12.63) | 78 vs. 81 | −3 | 0.70 |
|  | Psychosocial | 4.17 (−1.32, 9.65) | 80 vs. 76 | +4 | 0.14 | −3.18 (−12.80, 6.44) | 79 vs. 82 | −3 | 0.50 |
|  | Total | 4.32 (−1.39, 10.03) | 82 vs. 77 | +5 | 0.14 | −1.66 (−11.52, 8.20) | 81 vs. 82 | −1 | 0.73 |
| **Prednisone**  **(Yes vs. No)** | Physical | 0.77 (−6.91, 8.44) | 83 vs. 82 | +1 | 0.84 | 0.44 (−11.60, 12.47) | 83 vs. 83 | 0 | 0.94 |
|  | Emotional | 7.22 (1.63, 12.82) | 80 vs. 73 | +7 | **0.01** | 5.74 (−2.62, 14.10) | 77 vs. 71 | +6 | 0.17 |
|  | Social | 2.91 (−2.66, 8.48) | 88 vs. 85 | +3 | 0.30 | 8.13 (−0.83, 17.09) | 94 vs. 86 | −8 | 0.07 |
|  | School | 6.26 (−1.78, 14.30) | 76 vs. 69 | +7 | 0.13 | 9.26 (−3.13, 21.65) | 84 vs. 75 | +9 | 0.14 |
|  | Psychosocial | 5.70 (0.64, 10.76) | 81 vs. 76 | +5 | **0.03** | 7.78 (0.14, 15.42) | 84 vs. 77 | +7 | **0.05** |
|  | Total | 4.05 (−1.12, 9.21) | 82 vs. 78 | +4 | 0.12 | 5.49 (−2.39, 13.36) | 84 vs. 79 | +5 | 0.16 |
| **Azathioprine**  **(Yes vs. No)** | Physical | 3.93 (−3.68, 11.55) | 85 vs. 81 | +4 | 0.31 | 5.68 (−6.61, 17.98) | 86 vs. 80 | +6 | 0.35 |
|  | Emotional | 1.99 (−3.68, 7.66) | 76 vs. 74 | +2 | 0.49 | 0.43 (−8.14, 8.99) | 74 vs. 74 | 0 | 0.92 |
|  | Social | −0.49 (−6.03, 5.05) | 86 vs. 86 | 0 | 0.86 | 2.50 (−6.72, 11.73) | 91 vs. 89 | +2 | 0.58 |
|  | School | 3.66 (−4.22, 11.53) | 73 vs. 70 | +3 | 0.36 | 8.88 (−3.84, 21.61) | 84 vs. 75 | +9 | 0.16 |
|  | Psychosocial | 1.66 (−3.50, 6.81) | 78 vs. 77 | +1 | 0.53 | 3.14 (−4.72, 10.99) | 82 vs. 79 | +3 | 0.42 |
|  | Total | 2.49 (−2.81, 7.79) | 80 vs. 78 | +2 | 0.35 | 4.07 (−4.00, 12.14) | 83 vs. 79 | +4 | 0.31 |
| **Fatigue**  **(Yes vs. No)** | Physical | −16.69 (−25.30, −8.08) | 73 vs. 90 | −17 | **0.0003** | −13.37 (−23.86, −2.88) | 76 vs. 89 | −13 | **0.01** |
|  | Emotional | −11.22 (−17.36, −5.08) | 68 vs. 79 | −11 | **0.001** | −9.37 (−16.66, −2.08) | 70 vs. 79 | −9 | **0.01** |
|  | Social | −10.05 (−16.89, −3.20) | 81 vs. 91 | −10 | **0.005** | −7.86 (−15.68, −0.04) | 86 vs. 94 | −8 | **0.05** |
|  | School | −16.89 (−25.78, −7.99) | 63 vs. 80 | −17 | **0.004** | −13.28 (−24.10, −2.47) | 73 vs. 86 | −13 | **0.02** |
|  | Psychosocial | −12.71 (−18.44, −6.97) | 70 vs. 83 | −13 | **˂0.0001** | −10.29 (−16.96, −3.62) | 75 vs. 86 | −11 | **0.004** |
|  | Total | −13.88 (−19.69, −8.06) | 72 vs. 85 | −13 | **˂0.0001** | −11.13 (−18.00, −4.26) | 76 vs. 87 | −11 | **0.003** |
| **Pruritus**  **(Yes vs. No)** | Physical | −9.39 (−19.96, 1.17) | 76 vs. 85 | −9 | 0.08 | −3.96 (−17.23, 9.31) | 81 vs. 85 | −4 | 0.55 |
|  | Emotional | −6.29 (−13.97, 1.39) | 69 vs. 76 | −7 | 0.11 | −5.53 (−14.73, 3.68) | 71 vs. 77 | −6 | 0.23 |
|  | Social | −5.47 (−13.45, 2.52) | 82 vs. 87 | −5 | 0.18 | −5.86 (−15.72, 3.99) | 87 vs. 93 | −6 | 0.23 |
|  | School | −6.14 (−17.32, 5.03) | 68 v. 74 | −6 | 0.27 | −3.21 (−16.85, 10.44) | 78 vs. 81 | −3 | 0.63 |
|  | Psychosocial | −6.37 (−13.53, 0.79) | 72 vs. 79 | −7 | 0.08 | −5.27 (−13.67, 3.14) | 78 vs. 83 | −5 | 0.21 |
|  | Total | −7.09 (−14.25, 0.07) | 74 vs. 81 | −7 | **0.05** | -4.84 (-13.51, 3.83) | 79 vs. 84 | −5 | 0.26 |

^1^LSMean Diff is the difference between group-adjusted means (Yes – No).
